# Spreading Analysis of COVID-19 Epidemic in Bangladesh by Dynamical Mathematical Modelling

**DOI:** 10.1101/2020.06.12.20130047

**Authors:** A Fargana, A Arifutzzaman, A A Rakhimov

**Affiliations:** Department of Manufacturing and Material Engineering, International Islamic University Malaysia (IIUM), Kuala Lumpur, Malaysia; Research Centre for Nanomaterials and Energy Technology (RCNMET), School of Science and Technology, Sunway University, No. 5, Jalan Universiti, Bandar Sunway, Petaling Jaya, 47500 Selangor Darul Ehsan, Malaysia; Department of Science in Engineering, International Islamic University Malaysia (IIUM), Kuala Lumpur, Malaysia

**Keywords:** COVID-19, Modified SIR Model, Infection rate, Death rate, Growth rate, Basic Reproduction Number, Lockdown effect, Bangladesh

## Abstract

The coronavirus disease 2019 (COVID-19), which emerged from Wuhan, China, is now a pandemic, affecting across the globe. Bangladesh also is experiencing the rapid growth of COVID-19 infection and death cases started from 8^th^ March 2020. The purpose of providing a simple yet effective explanatory model for prediction of the future evolution of the contagion and verification of the effectiveness of the containment and lockdown measures in Bangladesh. In this study, using a modified SIR (Susceptible-Infected-Recovered) model a forecast is generated to predict the trends of COVID-19 cases in Bangladesh. The epidemic model was proposed to accommodate the effects of lockdown and individual based precautionary measures. Data has been taken and analyzed for before and after the movement control order (MCO) and during the MCO period. Modified SIR model in this work offers us an idea how the outbreak would progress based on the current data. It also has estimated that, the peak in terms of the number of infected cases will start from last of June 2020. For the total population (100%) the model gets the peaks at 214875 (infected cases) and 7743 (death cases). For the 90% population, the model shows the peaks at 244356 (infected cases) and 9100 (death cases). Analysis revealed that the lockdown and recommended individual hygiene can slow down the outbreak but unable to eradicate the disease from the society. With the current infection and death rate and existing level of personal precautionary the number of infected individuals will be increasing.

## 1. Introduction

A novel coronavirus, formerly called 2019-nCoV, or SARS-CoV-2 (Severe Acute Respiratory Syndrome Coronavirus 2) by ICTV (International Committee on Taxonomy of Viruses) caused an outbreak of atypical pneumonia, now officially called COVID-19 (Coronavirus Disease 2019) by World Health Organization (WHO) first in Wuhan, Hubei province in December 2019 and then rapidly spread out in the whole China as well as most of the parts of the world (Huang et al., 2020). Intermediated with the aid of huge aviation industry, it become into a world pandemic within only two months (Salim et al., 2020). As of June 11, 2020, the number of cases climbed above 7.58 million with a death toll of over 423,086 worldwide (JHU, 2020). The global impact and the public health threat of COVID-19 is the most serious seen in a respiratory virus since the 1918 influenza pandemic (Ferguson et al., 2020). Both COVID-19 and the 1918 influenza pandemic are associated with respiratory spread, a significant percentage of infected people with asymptomatic cases transmitting infection to others and a high fatality rate (Morens, 2018).

At 8 March 2020, first three coronavirus classes were detected and announced by the Institute of Epidemiology Disease Control and Research (IEDCR) in Bangladesh among around 111 tests. The cases included two men and one woman, who were aged between 20 to 35 years. Of them, two men were Italy returnees and the woman were a family member of one of these two. On March 16, the country detected three more cases of COVID-19, bring the total number of infected cases to eight. Bangladesh recorded first death a 70-year-old man due to COVID-19 on March 18. For slowing down the spreading in the country, authority tried to adopt numerous measures including growing awareness, compulsory lockdowns, home quarantine, social distancing and local or international flight bans among others. Bangladesh followed shutting down schools and colleges on March 18 and one week later from March 26 the all offices remain close resulting national lockdown (IEDCR, 2020). By 6^th^ of June 2020, 384851 tests had been performed as the disease spread to 64 districts and the country counted 63026 cases and a death toll of 846 persons (Bangladesh Government Press Releases, 2020).

The rapidly increasing demand on health facilities and health care workers threatens to leave some health systems overstretched and unable to operate effectively. In this situation it is very essential to have an accurate prediction of new cases due to COVID19 so that the necessary preparation by hospitals and requisite actions by administration can be taken in advance. Further, a necessary course of action is also needed to plan so that the country can tackle the situation as well as hurdle to spike to the next upper stages. The dire urgency in controlling the outbreak to prevent the collapse of healthcare system has forced the government to impose a more stringent action such as Movement Control Order (MCO).

During this anti-epidemic battle, besides medical and biological research, theoretical studies based on either statistics or mathematical modeling may also play a non-negligible role in understanding the epidemic characteristics of the outbreak, in forecasting the inflection point, ending time and in deciding the measures to curb the spreading. Several kinds of models have been proposed for describing the time evolution of epidemics, among which we distinguish two main groups: collective models and networked models. Susceptible-Infected-Recovered (SIR) models belong to the class of the so-called compartmental models (Brauer, 2017). Previously authors developed a infected disease model to predict the COVID-19 cases in Malaysia (Fargana and Arifutzzaman, 2020).

In this study, a mathematical model has been proposed for analysis of the scenarios of the community spreading of COVID-19 in the Bangladesh. An approximate prediction for the spread of COVID-19 in coming days could be achieved by using this model. This model can also be implemented for other countries as well to predict the number of COVID-19 cases in coming days. The proposed model will be able to predict the stages of the COVID-19 by comparing the available data with analytical results. Additionally, the other statistical analysis such as number of deaths or recovered cases, basic reproduction number and lockdown effect can also be estimated with the enough precision.

## 2. Data Collections

The numerical data of daily and total infections, fatalities and recovered was collected from the Bangladesh government’s source: Institute of Epidemiology and Disease Control and Research (IEDCR 2020) (www.iedcr.gov.bd). Total population of Bangladesh is collected from Wikipedia (www.wikipedia.org.wiki.Bangladesh). Movement control Order (MCO) phases (MCO1: 26^th^ March to 4^th^ April, MCO2: 4^th^ to 14^th^ April, MCO3:14^th^ to 25^th^ April, MCO4: 25^th^ April to 5^th^ May, MCO5: 5^th^ to 16^th^ May, MCO6: 16^th^ to 30^th^ May 2020) were collected from a daily newspaper named “The Daily Nayadiganto” (www.dailynayadiganta.com). On the other hand, presented world map with the coronavirus spots was collected from John Hopkins University (JHU, 2020) website shown in Figure 1.

**Figure 1:**
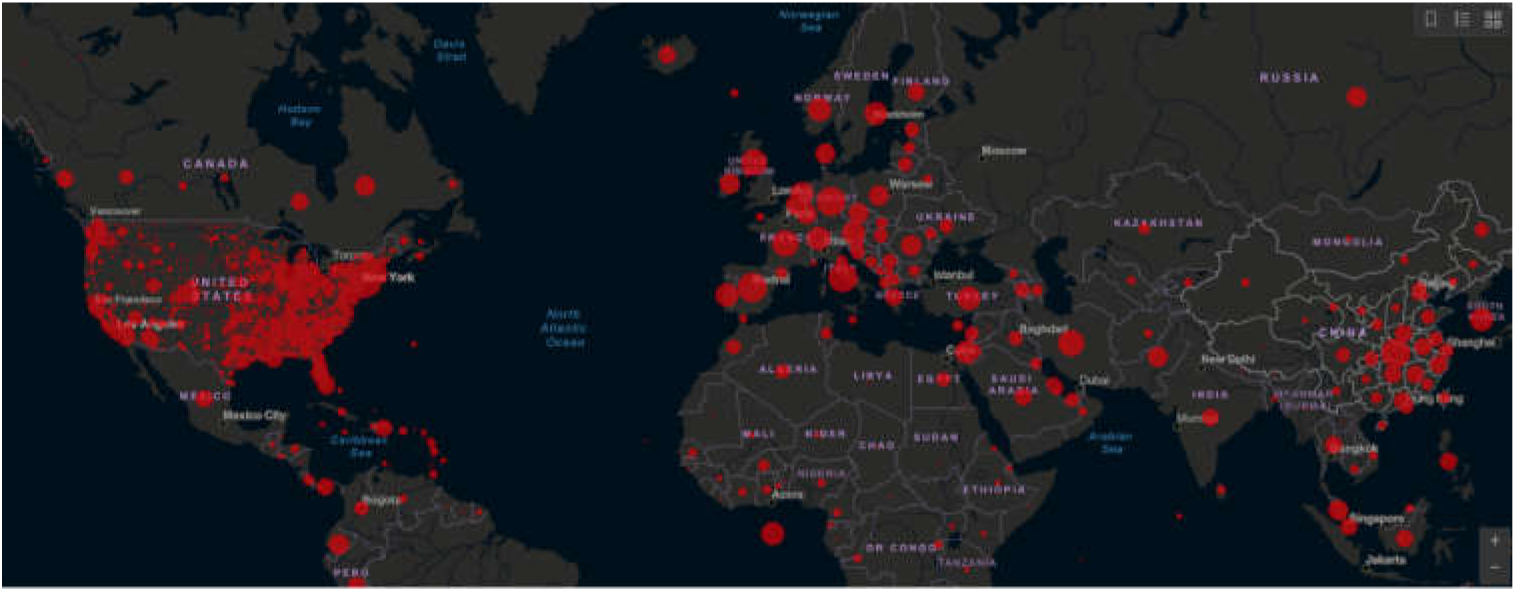
Coronavirus COVID-19 Global Cases by John Hopkins University (JHU, 2020) (www.coronavirus.jhu.edu.map.html).

## 3. Model

### 3.1 Modified Sir Model for Covid-19 Contagion

As SIR models discussed a compartmental disease model which includes three different compartments: ‘Susceptible’, ‘Infectious’ and ‘Recovered’. The population (N) is assigned as the sum of these three compartments. Susceptible individuals can become infected through contact with the infectious person and they have no immunity to the disease. Infectious people have already the disease or the symptoms and they can spread it to other peoples. Also, by recovering from the illness, infectious peoples can easily move into the ‘Recovered’ compartment. Finally, recovered peoples have the immunity from a prior exposure so that they can no longer become infected.

In SIR model, the number of individuals in each compartment can changes over time because individuals can move between the compartments. Therefore, SIR model captures all individuals’ changes in each compartment with ordinary differential equations (ODEs) to model the development of the COVID-19 pandemic. Figure 2 illustrates the schematic diagram of the basics of SIR model.

**Figure 2:**
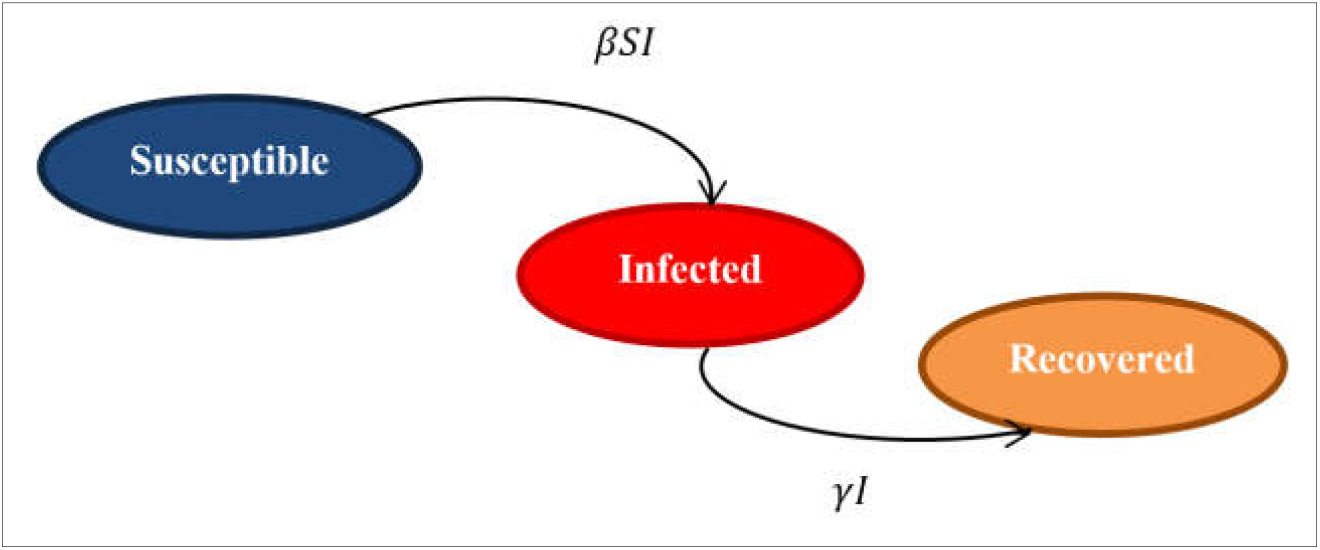
Schematic of the basics of SIR Model.

There are two parameters β and γ; where β is the rate of infection which means the rate of susceptible population get infected and γ is the death rate which means rate of infected population becoming die. Equations (1-3) describe the modified SIR model,

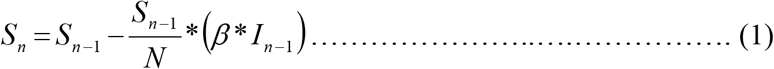

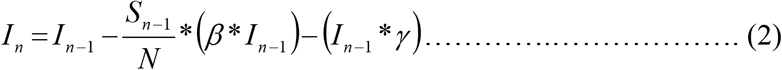

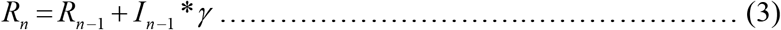

Equations (1-3) represent the rates of the components in SIR model. Notably, the natural death or birth rate is not considered in this model. Equation 1, 2 and 3 describe the susceptible (S), Infected (I), and death (R) people respectively, where *S*_*n*_, *I*_*n*_ and *R*_*n*_ represent the number people today, on the other hand *S*_*n* −1_, *I*_*n* −1_ and *R*_*n* −1_ represent the number of people yesterday. In Equation 1, the number of susceptible people will be reduced eventually because the individual gets infected gradually. In Equation 2, the infected persons are increasing with the infection rate, *β*. At the same time, the infected number of persons will be decreased because of the death rate *γ*. Basic Reproduction Number, *R*_0_ is the ratio of infection rate, *β* and the death rate, *γ*. The term *R*_0_ can be explained, if *R*_0_ < 1, then the pandemic is expected to decrease. If *R*_0_ =1, then the pandemic is expected to be stabilized which means number of infections remain same. If increased and infect higher number of populations. *R*_0_ > 1, then the pandemic will be

## 4. Results and Discussion

### 4.1. Current COVID-19 Statistics in Bangladesh

Until 6 June 2020, Bangladesh recorded total number of infected, recovered and death cases of 63026, 13903 and 846 respectively. Figure 3 represents the overview of the Covid-19 infected, recovered and death cases of Bangladesh from 8 March to 6 June, while Figure 4 represents the cumulative cases within this period.

**Figure 3:**
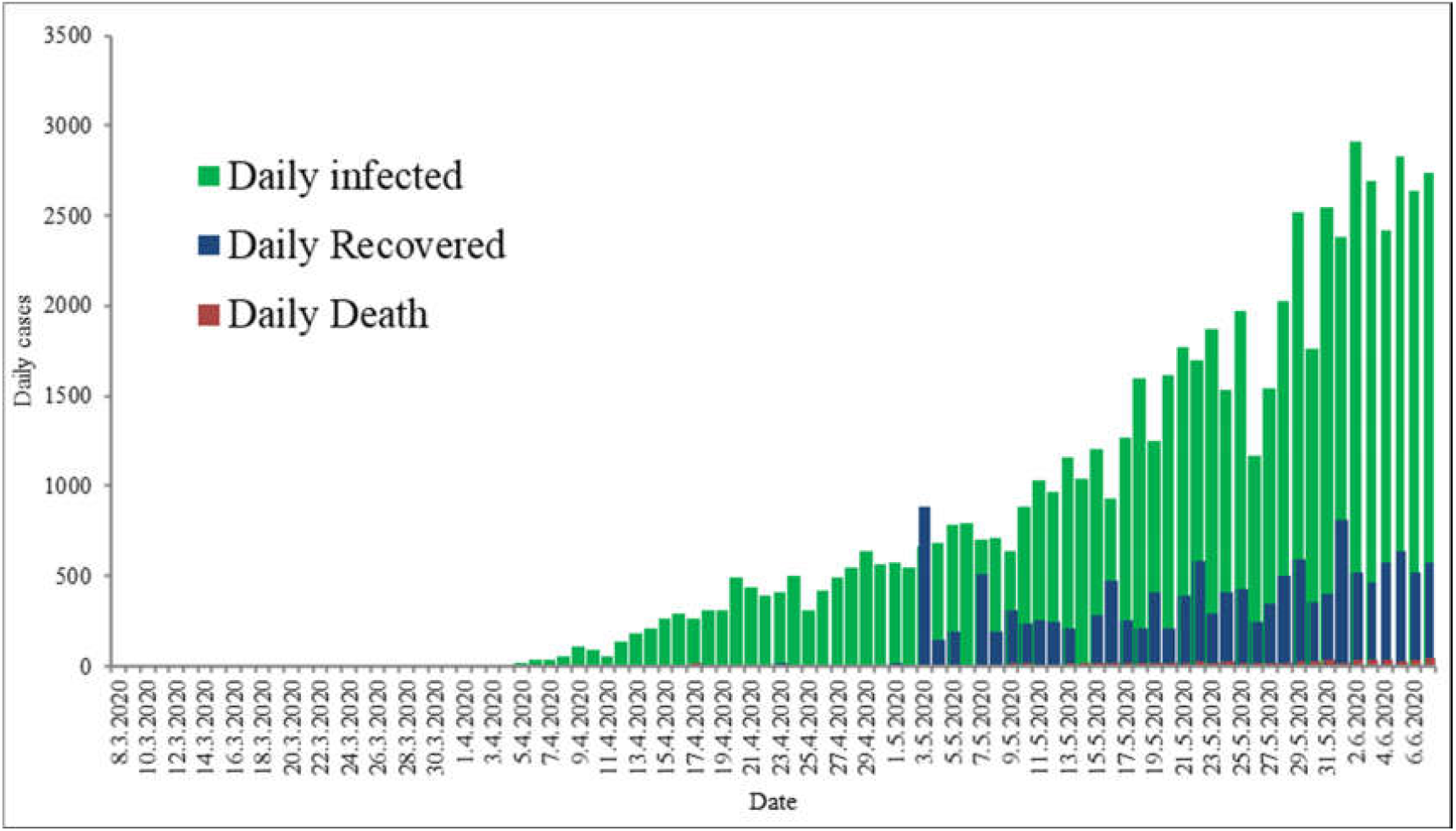
Recorded number of daily infected (green), recovered (blue) and death (red) cases in Bangladesh (8^th^ March to 6^th^ June 2020).

**Figure 4:**
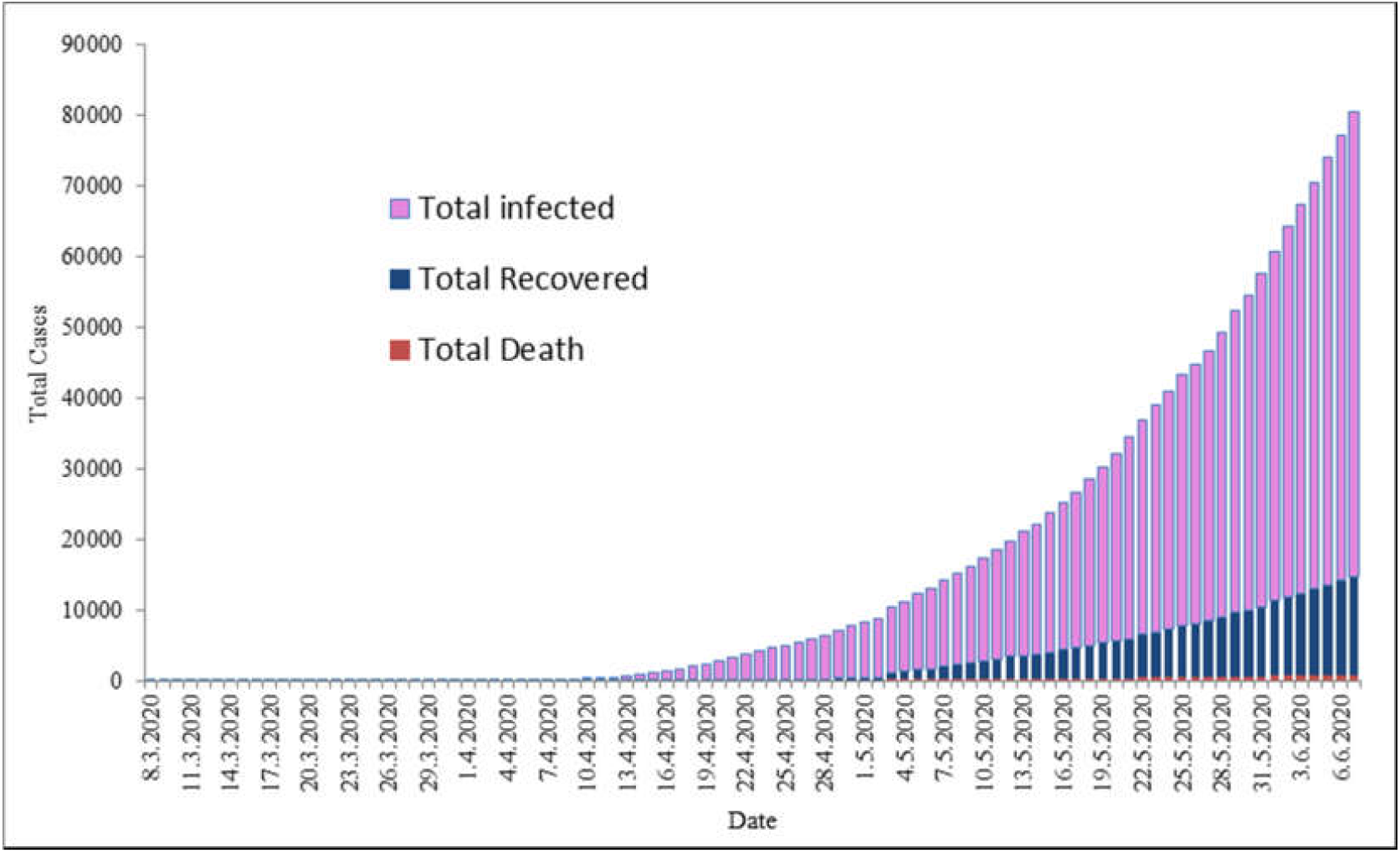
Cumulative infected (purple), recovered (blue), and death (red) cases in Bangladesh (8^th^ March to 6^th^ June 2020).

Figure 5 illustrates the infection growth rate in different MCO phases in Bangladesh. Collected data depicts that, the number of infected cases in Bangladesh is rising exponentially. In terms of the growth rate of the infected cases data is considered and analyzed from 8^th^ March to 6^th^ June 2020. It is seen that, the infection rate of COVID-19 in Bangladesh exposing the sign of increasing trend for the last few days with the average growth rate of ∼ 0.12. It is seen that, after detection of first COVID-19 case in Bangladesh within few days, administration took the necessary measures such as MCO. For this reason, before the implementation of the MCO the growth rate was not spikes drastically (before MCO the total case was only 39). It is detected that, the effect of MCO1and MCO2 was very marginal in controlling the contact among the infected persons which leads to the necessity of imposing further MCO. Until the MCO2 the growth rates found to be fluctuated after that, the difference of the growth rates was almost similar.

**Figure 5:**
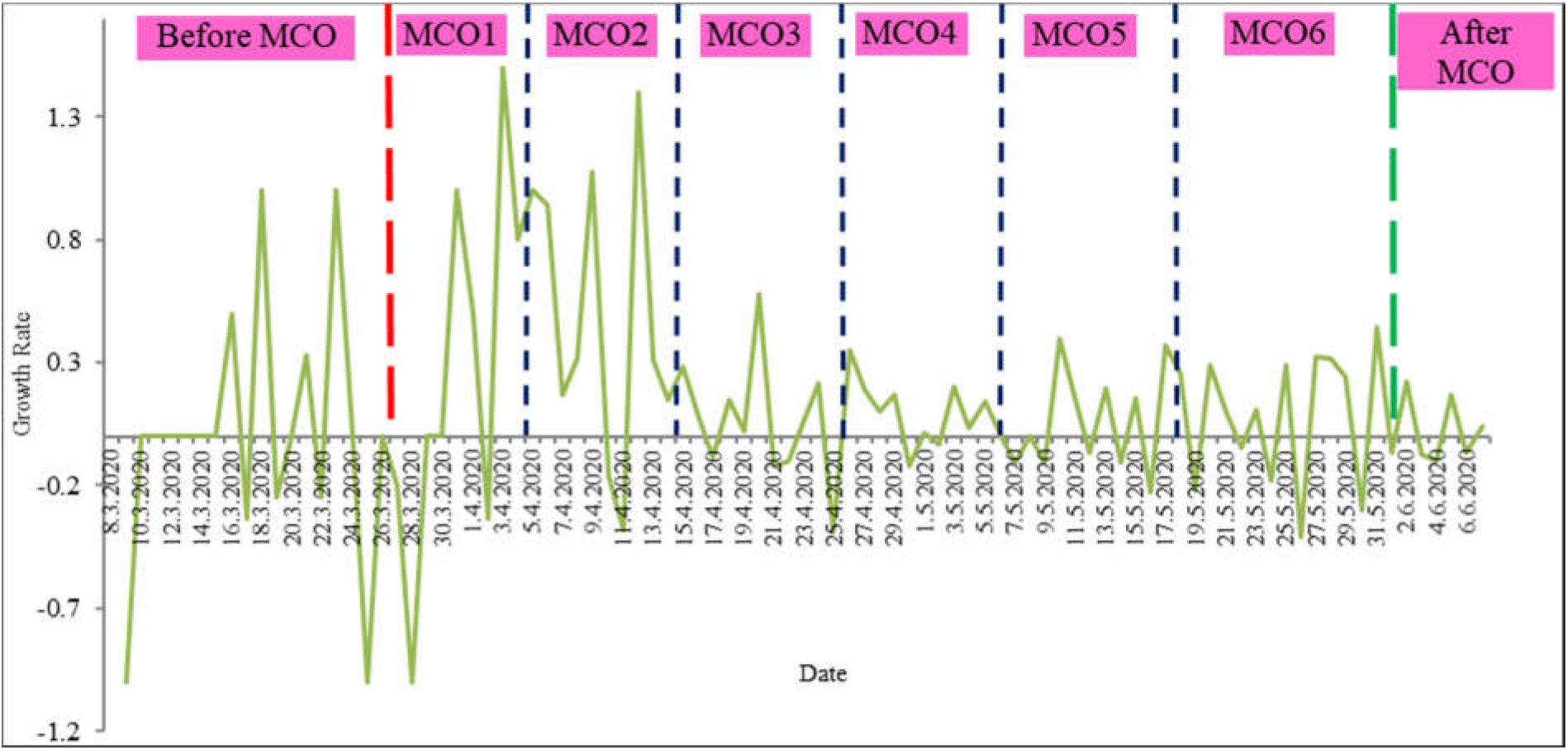
Growth rates of the COVID-19 infected cases in Bangladesh.

Therefore, relying on the strict social distancing method alone (like MCO) will not be the adequate measure to control the outbreak. Other necessary basic steps must be implemented such as identification of infected person. Because asymptomatic or symptomatic infected persons are easier to isolate from the other susceptible persons. Widespread and speedy contact tracing are desirable to identify all exposed person and put under quarantine to stop them from infecting other susceptible persons. Appropriate surveillance method is required to ensure compliance. If all these measures are not strengthened, MCO alone will not be enough even though it is accomplished with full force. Nonetheless, MCO is vital to decrease the immense contamination among the peoples so that the healthcare system gained adequate time for preparation.

Figure 6 shows the infection fatality rate (IFR) and cumulative fatality rate (CFR) of COVID-19 pandemic in Bangladesh for the period analyzed. IFR is the ratio of the confirmed death and infection cases of COVID-19 pandemic. Until, 6^th^ June 2020, the cumulative fatality rate of the COVID-19 in Bangladesh is detected about 4.08 % with the highest daily infection fatality rate of 33.33 % at 1^st^ April 2020.

**Figure 6:**
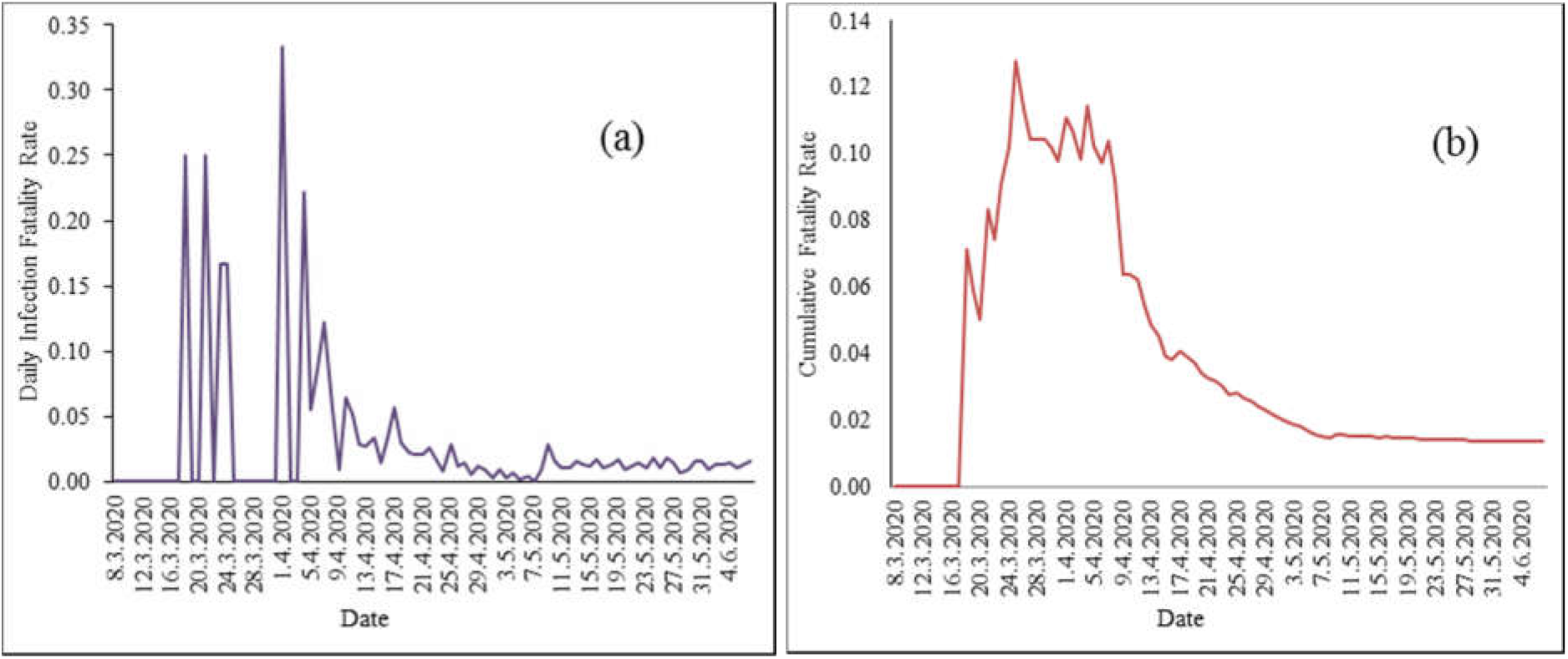
(a) Daily infection fatality rate and (b) cumulative fatality rate of COVID19 in Bangladesh (as of 6 June 2020).

### 4.2 Estimated Outcome of Modified SIR Model

Within the period considered (8^th^ March to 6^th^ June 2020) the infection rate, *β* and the death rate, *γ* of the COVID-19 patients were estimated for the two phase of population in Bangladesh. The total population of Bangladesh was considered 161 million. Two different population size was considered in the modified model for the analysis. First population size was 100 % and second population size was considered 90 %. As some people fully complied the order decreed by the administration which was considered about 10 %. This amount was not taken into account in second population size thought to be un-susceptible for the COVID-19 infection. At first, modified SIR model was implemented to find the known infection and death individual. It provides the error or appropriateness of the model. Figure 7 shows the estimated and actual infection and death individual of the total population (161 million). It was found that the error was ∼18 % between the estimated and actual infection individual. On the other hand, the error between the estimated and actual death was ∼ 23 %. These errors can be considered very minimal due to the such big population size (Salim et al., 2020). Thus, this model is found as appropriate for the future predictions. These errors will be taken into consider for the projected upcoming days.

**Figure 7:**
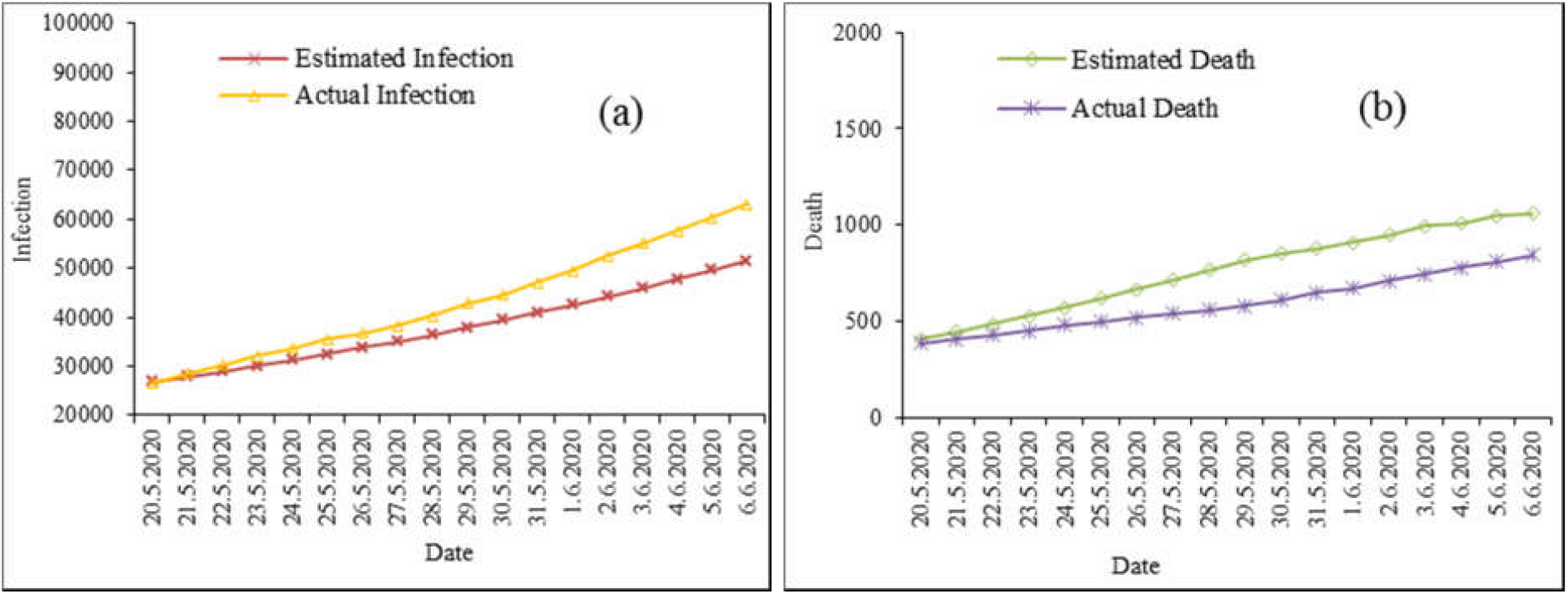
(a) Estimated infection and actual infection of total population and (b) Estimated death and actual death of total population of COVID-19 in Bangladesh (from 20 May 2020 to 6 June 2020).

Here, Figure 8 represents the predicted infected and death cases based on total population (100%). In general, this graph illustrates the impact of MCO. Therefore, the infected and death cases are still growing exponentially.

**Figure 8:**
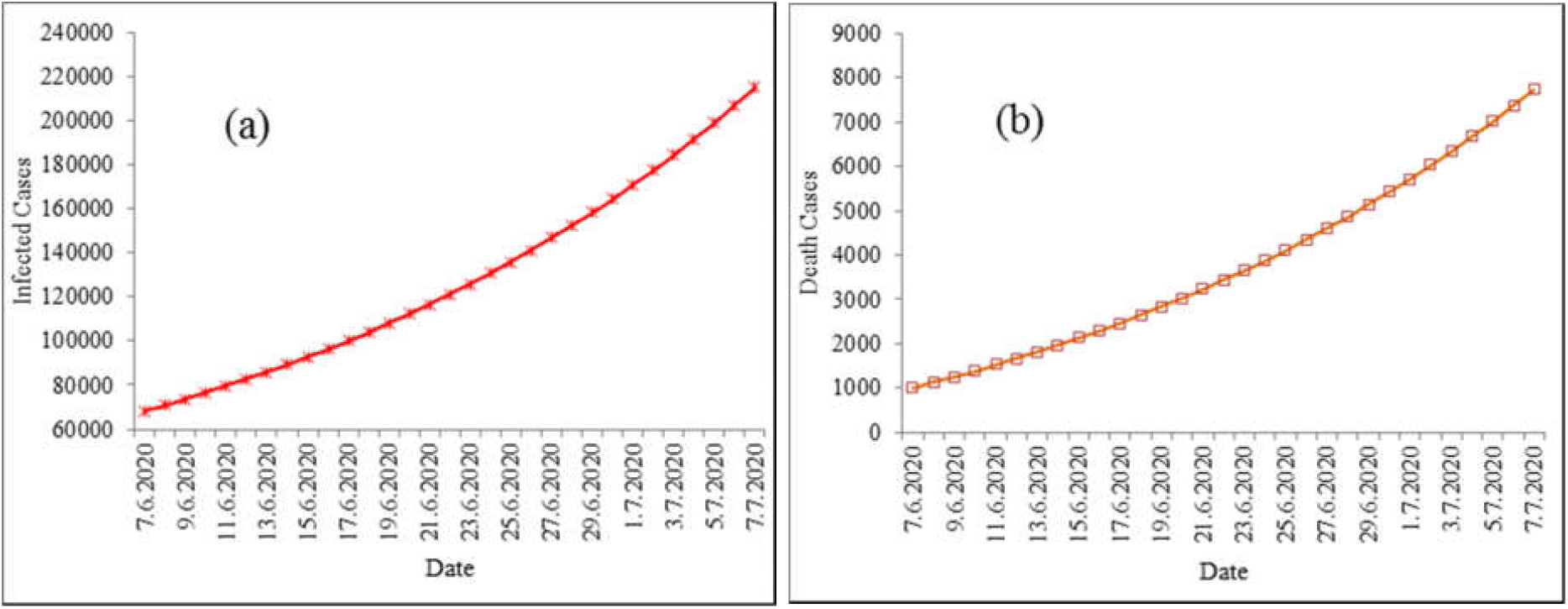
(a) Predicted infected people of total population and (b) Predicted death of total population in Bangladesh (from 7^th^ June 2020 to 7^th^ July 2020).

Figure 9 presents the predicted infected and death cases for the second population size (90 % population). This analysis also demonstrates the growing affected individuals. However, first analysis (out of 100% population) shows the less affected and death individual rather than the second analysis. A significant difference was perceived between the two population sizes (100 and 90%) analyzed. For the infected cases this difference was found ∼12.07% and for the death individuals it was ∼14.91 %.

**Figure 9:**
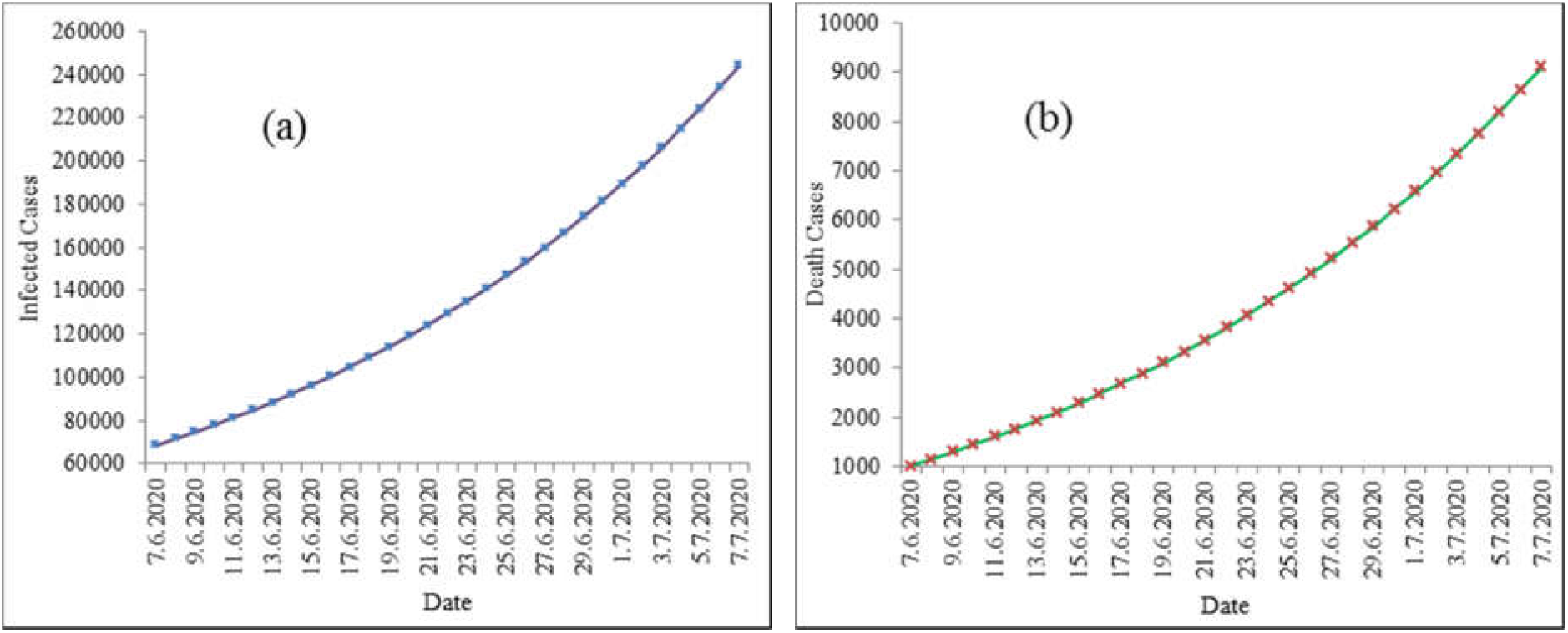
Predicted (a) infected cases and (b) deaths for the 90 % population in Bangladesh (7^th^ June to 7^th^ July 2020).

For the total population (100%) the model shows the peak for the infected number of ∼214875 and for the death cases of ∼7743 which are 0.13 and 0.0047 % of the total population of Bangladesh respectively. On the other hand, for the 90 % population the model peaks at 244356 (infected people) and 9100 (death people) which are about 0.15 and 0.0056 % of the total Bangladeshi population correspondingly. Table 1 summarizes the corresponding values for the both population size.

**Table 1:** Predicted maximum number of infected and death cases due to the on the estimated COVID-19 outbreak in Bangladesh (6 June to 7 July 2020).

| Method | Population | Infected Cases | Death Cases |
| --- | --- | --- | --- |
| Modified SIR Model | 100% | 214875 | 7743 |
|  | 90% | 244356 | 9100 |

It is seen that different population size provides the different predictions on the estimated COVID-19 peak dates and a sizeable variation in terms of the maximum number of people infected in Bangladesh.

Estimated infection rate (*β*), death rate (*γ*) and basic reproduction number (*R*_0_) are presented in Table 2. It is observed that, basic reproduction numbers are very high. It reveals that, the spreading of COVID-19 disease will increase and high number of populations will be infected in Bangladesh. To estimate the parameters the data is taken during the MCOs. So, from the basic reproduction number we can conclude that the population are not complying the government order to avoid or minimize contact with the coronavirus infected patients.

**Table 2.** Parameters generated by the modified SIR models based on 100% and 90% population of Bangladesh.

| Model | Population Size | $\beta$ | $\gamma$ | $R_0$ |
| --- | --- | --- | --- | --- |
| Modified SIR Model | 100% | 0.040754952 | 0.001789602 | 22.77319945 |
|  | 90% | 0.04528328 | 0.001988446 | 22.7732 |

Figure 10 represents the total infected cases vs number of days for both the total and 90% population of Bangladesh (7^th^ June to 7^th^ July 2020). It depicts that, for the total population, for crossing the first 50000 infected individual needed 17 days, second 50000 took 11 days and next 50000 took only 8 days. On the other hand, for 90% population for crossing first 50000 infected individual needed 16 days, second 50000 took 10 days and next 50000 took 7 days only.

**Figure 10:**
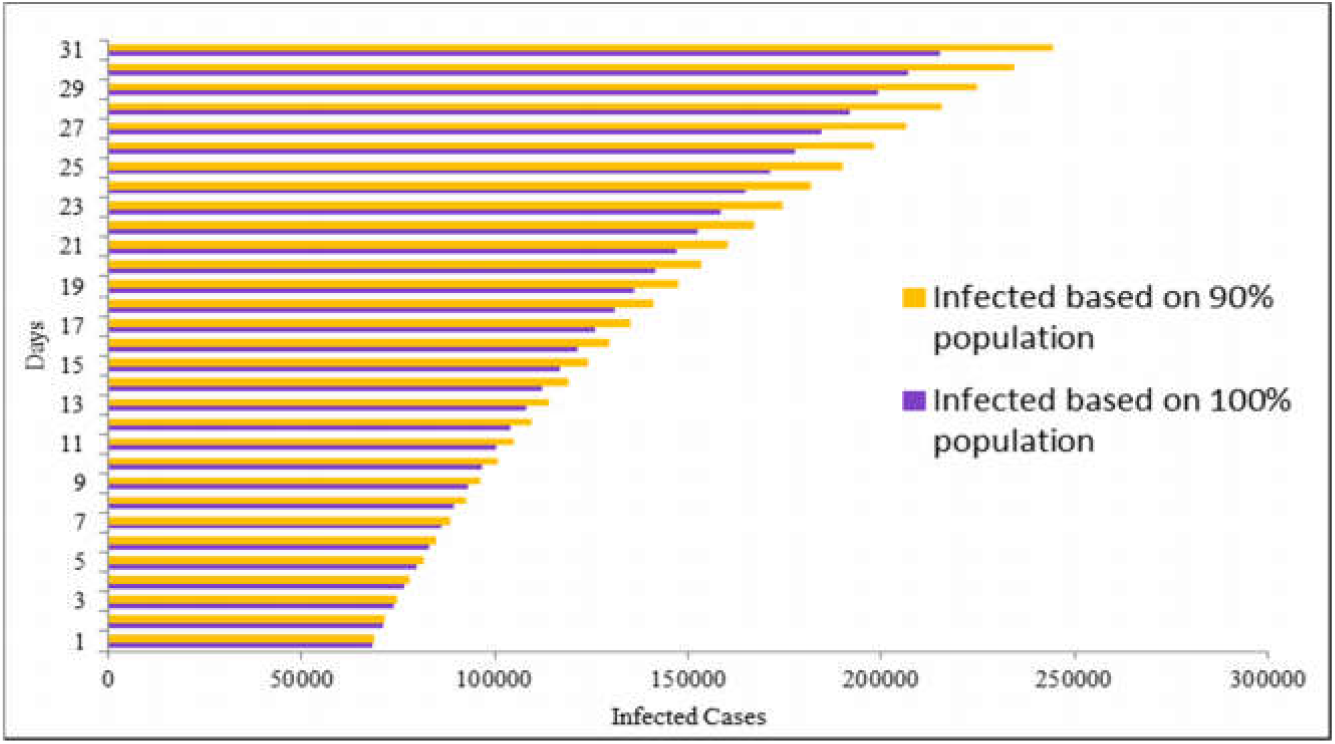
Total infected cases vs number of days for both the total and 90% population of Bangladesh (7^th^ June to 7^th^ July 2020).

## 5. Conclusion

Movement or restricted control order is crucial to mitigate the total number of people infected by COVID-19 and to ensure the health facilities can cope with the number at any given time. In this paper a forecast has been successfully generated using a modified SIR model to predict the trends of COVID-19 cases in Bangladesh. Modified SIR model in this work offers us an idea how the outbreak would progress based on the current data. It also has estimated that, the peak in terms of the number of infected cases will start from last of June 2020. It was found that, for the total population (100%) the model gets the peaks at 214875 (infected people) and 7743 (death people). For the 90% population, the model shows the peaks at 244356 (infected people) and 9100 (death people). Accurate prediction on the rising and declining period of COVID-19 cases could support the decision for MCO period and the expected level of compliance during the MCO periods. By analyzing the patterns and trends of the cases, forthcoming measures can be proposed and implemented. In addition, Bangladesh are still not doing extensive screening, as tests were only run on individuals that fulfill a specific criterion of potential COVID-19 and contacts of infected patients.

## Data Availability

In the reference list all the links have been provided from where data were collected.

